# Agricultural Injury Severity Prediction Using Integrated Data-Driven Analysis: Global Versus Local Explainability Using SHAP

**DOI:** 10.1101/2025.11.24.25340901

**Authors:** Omer Mermer, Yanan Liu, Charles A Jennissen, Milan Sonka, Ibrahim Demir

**Affiliations:** ByWater Institute, Tulane University, New Orleans, LA, USA; Iowa Initiative for Artificial Intelligence, University of Iowa, Iowa City, IA, United States; Roy J. and Lucille A. Carver College of Medicine, University of Iowa, Iowa City, IA, USA; Electrical and Computer Engineering, University of Iowa, Iowa City, IA, United States; River-Coastal Science and Engineering, Tulane University, New Orleans, LA, USA

**Author notes:** This manuscript is an medRxiv preprint and has been submitted for possible publication in a peer reviewed journal. Please note that this has not been peer-reviewed before and is currently undergoing peer review for the first time. Subsequent versions of this manuscript may have slightly different content.

**Keywords:** Agriculture, ensemble model, explainable artificial intelligence, injury severity prediction, machine learning, occupation

## Abstract

Despite the agricultural sector’s consistently high injury rates, formal reporting is often limited, leading to sparse national datasets that hinder effective safety interventions. To address this, our study introduces a comprehensive framework leveraging advanced ensemble machine learning (ML) models to predict and interpret the severity of agricultural injuries. We use a unique, manually curated dataset of over 2,400 agricultural incidents from AgInjuryNews, a public repository of news reports detailing incidents across the United States. We evaluated six ensemble models, including Gradient Boosting, XGBoost, LightGBM, AdaBoost, HistGradientBoosting, and Random Forest, for their accuracy in classifying injury outcomes as fatal or non-fatal. A key contribution of our work is the novel integration of explainable artificial intelligence (XAI), specifically SHapley Additive exPlanations (SHAP), to overcome the “black-box” nature of complex ensemble models. The models demonstrated strong predictive performance, with most achieving an accuracy of approximately 0.71 and an F1-score of 0.81. Through global SHAP analysis, we identified key factors influencing injury severity across the dataset, such as the presence of helmet use, victim age, and the type of injury agent. Additionally, our application of local SHAP analysis revealed how specific variables like location and victim’s role can have varying impacts depending on the context of the incident. These findings provide actionable, context-aware insights for developing targeted policy and safety interventions for a range of stakeholders, from first responders to policymakers, offering a powerful tool for a more proactive approach to agricultural safety.

## 1. Introduction

Occupational injury incidents impose significant burdens on individuals, organizations, and society at large, leading to considerable human suffering and economic costs [1]. The International Labor Organization estimates that approximately 337 million occupational incidents occur globally each year [2], driven by a complex interplay of contributing factors [3]. Given the extensive human and financial losses associated with such injuries, researchers have long sought to better understand the factors influencing incident severity and to improve the accuracy of predictive models for future events [4].

Among all occupational sectors, agriculture remains one of the most hazardous. In the United States, the agricultural sector consistently ranks near the top in injury rates, with 23.5 injuries per 100,000 workers reported in 2022 [5]. To mitigate these injuries, stakeholders must have access to detailed data about the causes and consequences of agricultural incidents. However, formal reporting requirements for agricultural injuries are often limited, resulting in sparse and incomplete national datasets.

Surveillance data plays a critical role in understanding agricultural injury patterns and informing prevention strategies. Previous research in agricultural injuries has involved collecting data from multiple sources such as surveys, news articles, workers’ compensation records, and electronic health systems [6]. Yet, challenges persist in capturing nonfatal injuries, accessing comprehensive surveillance data, and evaluating the cost-effectiveness of different data sources. For example, manual coding of narrative reports in large-scale surveillance efforts such as the Survey of Occupational Injury and Illness (SOII) demands considerable human labor, approximately 25,000 person-hours annually to process around 300,000 incidents [7].

To address these limitations, AgInjuryNews was established as a publicly accessible repository of news articles and reports documenting agricultural injuries and fatalities [8]. Curated by the National Farm Medicine Center (NFMC) and launched through the National Children’s Center for Rural and Agricultural Health and Safety (NCCRAHS), AgInjuryNews has compiled over 4,500 reports covering more than 6,000 victims between 2015 and 2024 [9]. It employs keyword filtering and digital media monitoring to identify relevant incidents, which are then manually reviewed and summarized by expert volunteers [10, 11]. While the structured nature of the data supports research and policy analysis, the system’s dependence on manual curation limits both scalability and timeliness.

Predicting the severity of agricultural injuries is a critical area of study within occupational safety research. Understanding the factors that influence injury severity enables the design and implementation of proactive safety interventions. The severity of such incidents is often shaped by a combination of human, environmental, and situational variables [12]. Moreover, spatial variability in agricultural practices and environments necessitates localized analyses to accurately predict and mitigate risks.

Historically, injury severity modeling has relied on statistical approaches, particularly discrete choice models such as binary logit, multinomial logit, and probit [13, 14]. More advanced frameworks, including Bayesian hierarchical models, multivariate models, and random parameter models, have attempted to address unobserved heterogeneity and variable correlations [15–17]. While these approaches offer interpretable insights, they depend on strong assumptions about variable distributions and linear relationships, which can result in biased estimates if violated [18–20]. Moreover, multicollinearity, unobserved heterogeneity, and high computational costs often hinder their practical application [21, 22].

With the rise of machine learning (ML), new models have emerged that address many limitations of traditional approaches. ML models such as Artificial Neural Networks (ANN), Support Vector Machines (SVM), Naïve Bayes (NB), and Decision Trees (DT) have demonstrated strong capabilities in modeling complex, nonlinear relationships between predictors and outcomes in many domains [23–25], including agriculture [26]. For instance, Abdelwahab and Abdel-Aty [27] reported that multilayer perceptrons outperformed traditional models in crash severity prediction. SVMs have shown superior performance in diverse contexts, including diverging segments on Florida freeways [28] and work zones [29]. NB models, while simple, are effective in binary classification tasks [30, 31], and DTs are valued for their interpretability in injury classification [32, 33].

More recently, ensemble learning techniques such as Random Forests (RF) and eXtreme Gradient Boosting (XGB) have gained prominence due to their superior predictive power and robustness [34–36]. RF has demonstrated consistent superiority over both statistical models and individual ML algorithms in highway crash severity prediction [37, 38]. XGB has proven especially effective in real-time crash detection [39], automated vehicle crash analysis [40], and fatal crash classification [41], offering a balance between accuracy and computational efficiency.

Interpretability remains a central challenge in applying ML models to safety-critical domains [42]. Ensemble methods such as XGB offer high predictive accuracy but are often regarded as “black boxes,” while inherently interpretable models like linear regression or decision trees provide transparency but may lack flexibility for complex patterns [43]. Explainable artificial intelligence (XAI) has emerged to address this trade-off [44], and SHapley Additive exPlanations (SHAP) is widely adopted for both global and local interpretability by assigning contribution scores to each input feature [45]. SHAP has been successfully applied in injury severity modeling to enhance transparency while maintaining performance. Ensemble models with SHAP identified critical features such as vehicle count and road category in New Zealand [46] and highlighted key risk factors in two-wheeler crashes, including helmet use and urbanization levels [47].

Despite widespread use of XGB, Light Gradient Boosting Machine (LightGBM), and Categorical Boosting (CatBoost), few studies systematically compare these models on real-world crash datasets while evaluating both predictive performance and interpretability. SHAP has been used to examine contextual differences in risk profiles across rural and urban environments [48, 49], interactions among road geometry, speed, and weather [50], and dominant contributors such as alcohol use, speeding, and seatbelt noncompliance in multilayer perceptron models [51]. SHAP TreeExplainer has also generated ranked feature importance lists across diverse crash contexts [52]. Collectively, these studies show that integrating SHAP with ML models provides actionable insights and improves interpretability, addressing the crucial balance between predictive accuracy and transparency in traffic injury severity modeling.

Despite the increasing adoption of global SHAP analysis, the application of local SHAP explanations, designed to interpret individual predictions, remains limited in both traffic and agricultural injury severity modeling. This limitation constrains the practical utility of ML in operational settings, where stakeholders such as first responders, planners, and safety specialists require case-specific insights to support timely and context-aware decisions. Local SHAP visualizations, including force plots, provide detailed accounts of how individual features influence predictions, capturing both the direction and magnitude of their effects. Research in other fields has demonstrated the added value of local interpretability: in solar radiation forecasting, global-only analyses obscured critical localized insights [53]; in water quality modeling, local SHAP revealed site-specific influences of environmental and meteorological factors [54]; and in ophthalmology, local explanations enhanced individual prediction of visual field parameters from optical coherence tomography data [55].

Collectively, this evidence underscores the importance of extending local SHAP approaches to injury severity modeling, where instance-level interpretability is vital for translating machine learning predictions into actionable outcomes. Without local interpretability, findings remain generalized and less useful for informing incident-specific interventions. Notably, while ML methods have been extensively explored in traffic crash severity research, their application to agricultural injuries remains limited, especially within the U.S. Few studies have evaluated a broad range of ML algorithms in this domain or offered structured, interpretable analyses of the contributing factors.

This study addresses these gaps by introducing a comprehensive ensemble ML framework for predicting agricultural injury severity. Specifically, we employ advanced gradient boosting models, including Gradient Boosting (GB), XGB, LightGBM, Adaptive Boosting (AdaBoost), Histogram-based Gradient Boosted Regression Trees (HistGBRT), and RF, alongside explainable ML techniques. The novelty of our work lies in its dual focus on predictive accuracy and model interpretability, particularly through the use of both global and local SHAP analyses. Our key contributions include: (1) leveraging a structured dataset from AgInjuryNews to model agricultural injury severity outcomes; (2) evaluating and comparing multiple ensemble ML models in terms of accuracy and training efficiency; (3) predicting injury severity using real-world data from diverse agricultural incidents; and (4) identifying and interpreting key contributing factors through SHAP-based feature importance analysis, both globally and locally, to support policy-making and intervention strategies.

The remainder of this paper is structured as follows: Section 2 outlines the dataset, featured engineering processes used, and the machine learning models implemented in the study. Section 3 presents experimental results, including model evaluation metrics, and SHAP-based insights. Section 4 discusses the findings, and Section 5 concludes with key takeaways and directions for future research.

## 2. Methodology

### 2.1. Data Description

The dataset used in this study was obtained from AgInjuryNews.com, an open-access surveillance platform that aggregates news media reports of agricultural injuries and fatalities occurring across the United States and Canada [7, 8]. Covering a time span from 2016 to 2024, AgInjuryNews collects incident-level data that includes a wide range of variables such as geographic location, demographic information, incident characteristics, and the presence or absence of safety equipment. These records are identified through automated keyword filtering and digital media monitoring, after which expert volunteers manually review and code the information into a structured format [9, 11].

To prepare the data for this study, the raw dataset from AgInjuryNews.com, which initially contained reports from both the United States and Canada, was thoroughly cleaned. During the preprocessing, records with missing values and duplicate entries were excluded, and the scope was narrowed to incidents reported only within the United States. The resulting dataset was comprised of 2,451 valid records of agricultural injury incidents. Of these, 778 cases (32%) were classified as non-fatal, and 1,673 cases (68%) were identified as fatal. The binary target variable in this study, injury severity, was coded as 1 for fatal incidents (in which at least one person died) and 0 for non-fatal incidents (which included injuries of all severity and incidents with property damage only).

Figure 1 presents the annual distribution of injury severity categories—fatal and non-fatal—between 2016 and 2023. Fatal injuries consistently outnumbered non-fatal ones across all observed years in the dataset. The frequency of incidents peaked in 2018, followed by a gradual decline, especially in fatalities. In 2020, the proportion of non-fatal injuries increased to 33% of that year’s total incidents, despite an overall decrease in reported cases. At the time this project began, in early 2024, only about 30 incidents from that year had been entered into the AgInjuryNews database. Since there is often a lag in reporting and entering new cases, we used all available data for machine learning model development. However, for visualization and trend analysis purposes, we present only the data from 2016 to 2023, where reporting is more complete.

**Figure 1:**
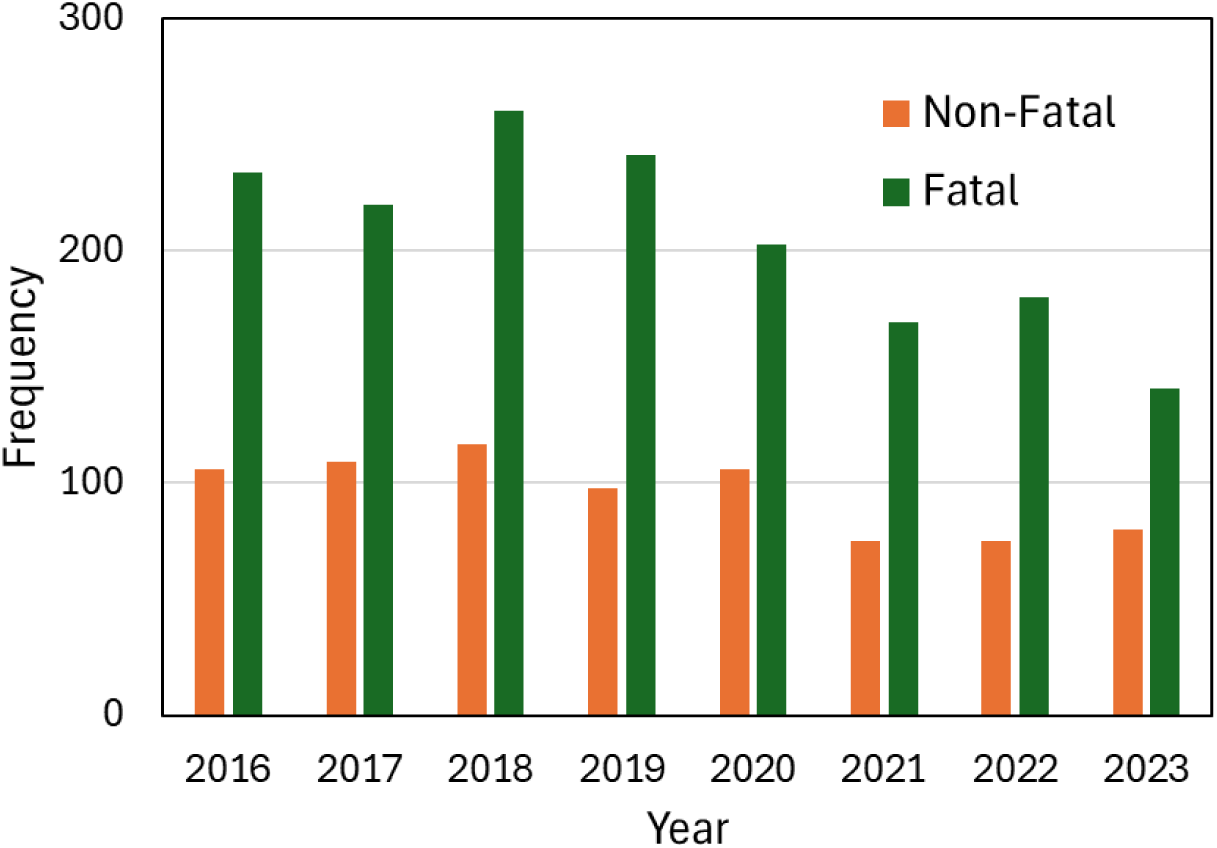
Annual distribution of agricultural injuries by severity from 2016 to 2023.

The modeling framework employed in this study utilized a comprehensive set of features that were grouped into four major categories: temporal attributes, personal characteristics, incident attributes, and environmental factors. These explanatory variables are summarized in Table 1 and include both categorical and continuous types, carefully selected based on their relevance to injury risk and severity. Temporal attributes included the season of occurrence (spring, summer, autumn, or winter), whether the incident occurred on a weekday or weekend, the time of day (morning, afternoon, evening, or night), and the calendar month. These variables allowed for the exploration of patterns related to seasonal work, time-of-day effects, and potential influences of agricultural cycles.

**Table 1.** Detailed description of all variables in the dataset.

|  | Variable Category | Features | Type | Description / Labeling |
| --- | --- | --- | --- | --- |
| Independent Variables | Temporal Attributes | Season | Categorical | 1: Spring (Mar-May); 2: Summer (Jun-Aug); 3: Autumn (Sep-Nov); 4: Winter (Dec-Feb) |
|  |  | Day | Binomial | 0: Weekday (Monday to Friday);<br>1: Weekend (Saturday and Sunday) |
|  |  | Time | Categorical | 1: Morning (6am – 12pm); 2: Afternoon (12pm – 6pm); 3: Evening (6pm – 12am);<br>4: Night (12am – 6am) |
|  |  | Month | Categorical | [1,12] |
|  | Personal Features | Gender | Binomial | 0: Male; 1: Female |
|  |  | Age | Continuous | [0, 98] |
|  |  | Alcohol | Binomial | 0: No; 1: Yes |
|  |  | Seatbelt | Binomial | 0: No; 1: Yes |
|  |  | Helmet | Binomial | 0: No; 1: Yes |
|  |  | Drowning / Suffocation | Binomial | 0: No; 1: Yes |
|  |  | ROPS | Binomial | 0: No; 1: Yes |
|  |  | Other PPE | Binomial | 0: No; 1: Yes |
|  | Incident Features | Injury Agent | Categorical | 0-2: ATV/Off-Road; 3-5: Building; 6-10: Env.; 11-12: Fall; 13-19: Fish/Forestry; 20-24: Livestock; 25-38: Machinery; 39-49: Others; 50-53: Pesticide; 54-59: Plant; 60-63: Skid Steer 64-69: Tractor; 70-76: Vehicle |
|  |  | # of Victims | Continuous | [1, 17] |
|  |  | Location | Categorical | 1: Roadways; 2: Agricultural Env.;<br>3: Forestry; 4: Fishing |
|  |  | Role | Categorical | 0: Bystander; 1: Certified First Responder;<br>2: Operator; 3: Other; 4: Passenger;<br>5: Worker/Farmer/Fisher |
|  |  | Intentional | Binomial | 0: No; 1: Yes |
|  |  | State | Categorical | [0, 49] |
|  | External Factors | Confined Space | Binomial | 0: No; 1: Yes |
|  |  | Grain Involved | Binomial | 0: No; 1: Yes |
|  |  | Agri-tourism | Binomial | 0: No; 1: Yes |
| Dependent Variable |  | Injury Severity | Binomial | 0: Fatal; 1: Non-Fatal |

Personal features included the victim’s gender, age, alcohol consumption, and the use of safety equipment such as seatbelts, helmets, and other personal protective equipment (PPE). Additional binary indicators like drowning and the presence of rollover protective structures (ROPS) were also included. These factors provide insight into the role of human behavior and safety compliance in injury outcomes. Incident features captured information about the agent involved in the injury—categorized into types such as machinery, livestock, ATVs, tractor or environmental hazards—as well as the total number of victims, the individual’s role (e.g., operator, bystander, passenger), and whether the incident was intentional. Geographic state was included to account for potential regional variations in practices or regulations.

External features were also incorporated, including whether the incident occurred in a confined space, whether grain was involved, and whether the location was part of an agritourism activity. These variables reflect the physical context of the incident and its potential contribution to severity. The outcome variable, injury severity, was binarized into fatal and non-fatal for supervised machine learning classification.

### 2.2. Machine Learning Models

In this study, we utilized advanced gradient boosting models, specifically Gradient Boosting (GB), XGBoost (XGB), LightGBM, AdaBoost, and HistGBRT, along with Random Forest (RF), and integrated explainable machine learning (ML) techniques.

**Gradient Boosting (GB):** The GB algorithm developed by Freund & Schapire [56], is a decision-tree based ensemble learning technique that utilizes the boosting method to improve accuracy. Boosting works through training each model in an ensemble individually and iteratively, where each new model attempts to correct the errors made by previous models. GB minimizes a loss function through the gradient descent method, where new trees or learners in a sequence are fitted to the residuals from the loss function with respect to the model’s current predictions [57]. This allows the model to achieve increased predictive performance through focusing on the most difficult-to-learn data points [58]. Additionally, the algorithm also takes advantage of regularization using parameters such as learning rate and maximum depth to help prevent overfitting through adding a penalty to a model’s complexity and improving robustness. These techniques result in a higher predictive performance for the gradient boosting algorithm, outperforming linear regression models and decision trees on non-linear, diverse, and complex datasets on both classification and regression tasks [59].

**Extreme Gradient Boosting (XGB):** XGB, introduced by Chen and Guestrin [60], is a widely used ensemble learning algorithm based on decision trees, designed to enhance both computational efficiency and model flexibility in supervised learning tasks. XGB implements a sophisticated form of the gradient boosting framework by iteratively combining the predictions of multiple weak learners—typically decision trees—into a single, robust model. At each iteration, the model updates the previous learner by correcting its errors, using gradient-based optimization guided by a defined objective function to assess improvements in performance [61]. To prevent overfitting and improve generalization, XGB integrates multiple explicit regularization techniques, including L1 (alpha), L2 (lambda), and structure-based penalties like gamma, into its objective function [62]. It also provides feature-important measures (e.g., gain, cover, frequency), which are particularly useful for interpreting models and managing high-dimensional data. The algorithm also employs a random sampling strategy to reduce variance and enhance the predictive power of the ensemble. Furthermore, XGB leverages both first- and second-order derivatives of the loss function, enabling more precise gradient direction estimation and thus more efficient minimization of the loss compared to traditional gradient boosting methods. Its support for parallel and distributed computing further contributes to reduced training times and more effective model exploration. XGB is highly efficient with large datasets and has been used in diverse fields from pedestrian injury severity prediction [63] to railroad incident analysis [64].

**Light Gradient Boosting Machine (LightGBM):** LightGBM framework, introduced in 2017, employs two advanced techniques, Exclusive Feature Bundling (EFB) and Gradient-based One-Side Sampling (GOSS), to efficiently handle large datasets with numerous features [65]. EFB reduces dimensionality by grouping mutually exclusive features, thereby minimizing unnecessary computations for sparse features [66]. LightGBM constructs trees leaf-by-leaf, addressing leaf imbalance, while GOSS selects split points based on variance gain rather than traditional information gain, enhancing efficiency in identifying optimal splits [67]. Additionally, LightGBM uses a histogram-based algorithm to discretize continuous feature values into k bins, aggregating statistics to determine segmentation scores. This approach balances speed and accuracy by replacing conventional pre-sorted methods [68] and also contributes to regularization, helping prevent overfitting.

**Adaptive Boosting (AdaBoost):** AdaBoost, introduced by Freund and Schapire [69], is the first and one of the most influential boosting methods. It operates on the principle that a strong classifier can be formed by sequentially combining multiple weak classifiers, each contributing to the final prediction. The algorithm employs a convex loss function and is known to be sensitive to noise and outliers in the dataset. AdaBoost adjusts the weights of training samples at each iteration, emphasizing misclassified instances to enhance the accuracy of subsequent weak classifiers [70]. This iterative refinement continues until a specified classification error threshold is achieved, effectively reducing bias in complex prediction tasks when combined with other boosting strategies [71]. Unlike some ensemble methods, AdaBoost is relatively resistant to both overfitting and underfitting in certain classification contexts. It improves classifier performance by updating the sample distribution and iteratively incorporating only the most effective classifiers while discarding the weakest ones [72]. This dynamic process leads to a cumulative increase in prediction accuracy. The detailed information and application of AdaBoost to investigate incident injury severity prediction can be found in various research articles [73–76].

**Histogram Gradient Boosted Regression Trees (HistGBRT):** HistGBRT is an advanced variant of the widely recognized gradient boosting ensemble method, commonly employed for both regression and classification tasks. Its primary goal is to enhance prediction accuracy by iteratively converting weak learners into a strong predictive model [77]. This is achieved through a sequential training approach, where each new weak learner is trained to correct the errors made by its predecessors [78]. HistGBRT specifically addresses key limitations of traditional gradient boosting methods, such as prolonged training times on large datasets. This issue is mitigated by discretizing continuous input features into a finite set of values, which accelerates the training process [79]. Unlike other boosting techniques, HistGBRT stores continuous feature values in specialized containers and uses these to construct histograms during training, leading to improved efficiency and reduced memory consumption [80].

**Random Forest (RF):** RF is a widely adopted machine learning algorithm known for its strong performance in both classification and regression tasks. Its effectiveness stems from the ensemble approach, where multiple randomized decision trees are constructed, and their outputs are aggregated, typically by averaging for regression or majority voting for classification. This methodology enhances predictive accuracy, particularly in high-dimensional settings where the number of variables exceeds the number of observations [81]. RF is also recognized for its flexibility, making it suitable for a wide array of learning tasks and scalable to large datasets. One of its key advantages is robustness to overfitting, achieved by averaging multiple de-correlated trees, which mitigates the risk of modeling noise from the training data [82]. Additionally, RF is resilient to outliers and can manage missing data without substantial performance loss. The algorithm provides interpretable metrics such as feature importance scores, which are valuable for understanding variable contributions. Recent advancements in crash severity prediction and related fields have prominently utilized RF [12, 37].

### 2.3. Model Development

The comprehensive ML workflow for agricultural injury severity prediction is illustrated in Figure 2. Following cleaning and preprocessing of AgInjuryNews dataset, fivefold cross validation, which is randomly partitioned into five equal sized subsamples, were applied to all ensemble models. This approach allows us to reduce bias by utilizing all data for training and testing, and a mitigation of overfitting by assessing model performance across multiple unseen data subsets. The target variable, injury severity, was treated as a binary classification problem (fatal vs. non-fatal). All models for this study were implemented using Python.

**Figure 2.**
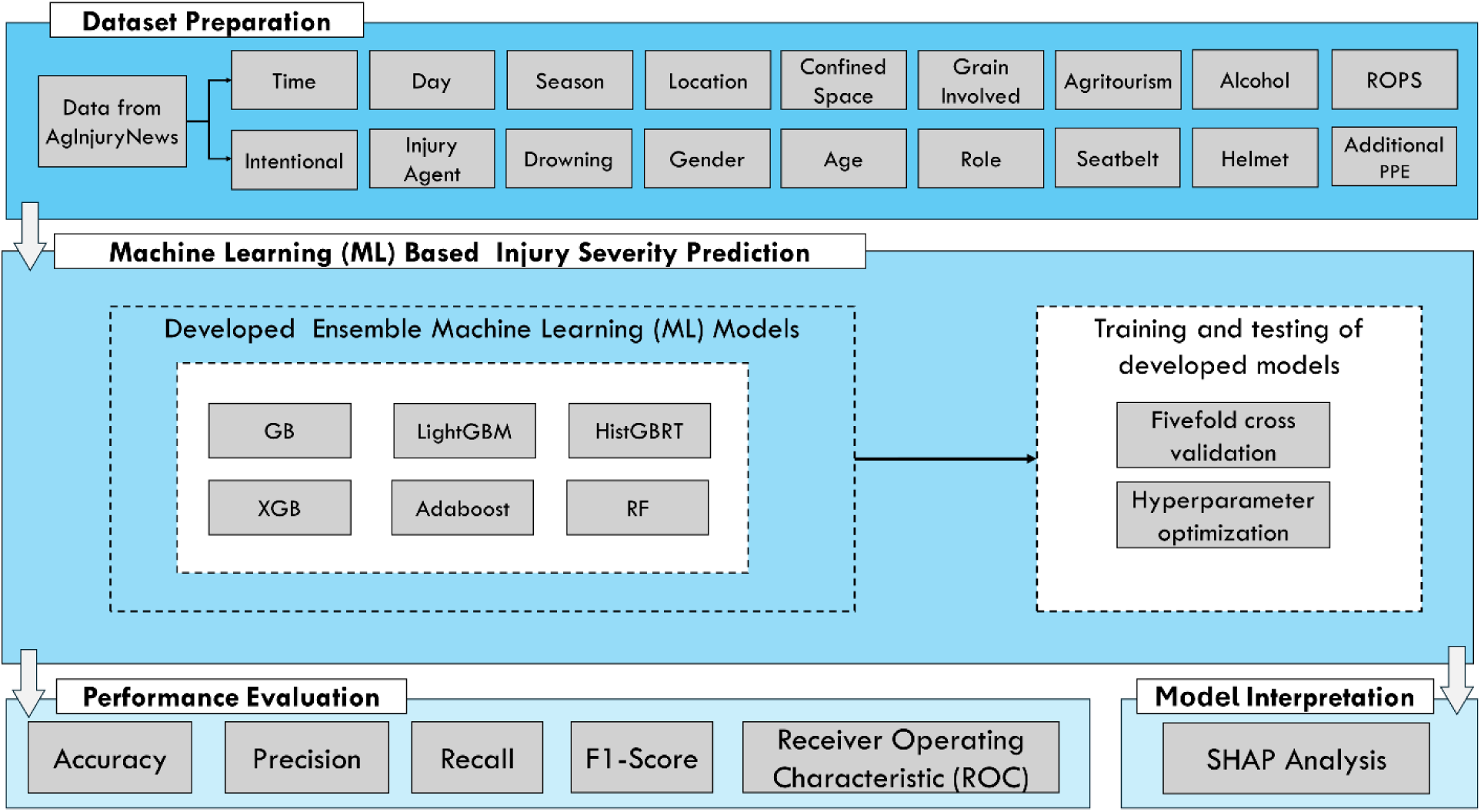
Workflow diagram for all scenarios in agricultural injury severity prediction.

A variety of ensemble models—including GB, XGB, LightGBM, AdaBoost, HistGRBT, and RF—were then trained and tested to predict agricultural injury severity. The hyperparameters for each model were systematically tuned using cross-validation, a robust technique that partitions the training data into multiple subsets. This allowed us to evaluate model performance under different parameter settings, ultimately identifying and selecting the most accurate configuration for each algorithm. This optimization process was critical for maximizing the predictive performance of the models and ensuring their robustness.

### 2.4. Model Evaluation

When comparing classification models, various performance metrics derived from a confusion matrix are commonly used. As shown in Table 2, a confusion matrix is a contingency table that visualizes the performance of an algorithm by illustrating how observations are distributed across actual and predicted classes. The matrix comprises four key outcomes: True Positives (TP), where the model correctly predicts the positive class; True Negatives (TN), where it correctly predicts the negative class; False Positives (FP), where it incorrectly predicts the positive class; and False Negatives (FN), where it incorrectly predicts the negative class. This matrix provides a clear and comprehensive representation of a model’s classification accuracy and errors.

**Table 2.** Confusion matrix for binary classification.

| Table 2. Confusion matrix for binary classification |  |  |
| --- | --- | --- |
| Actual Class | Predicted Class |  |
|  | Positive | Negative |
| Positive | True Positive (TP) | False Negative (FN) |
| Negative | False Positive (FP) | True Negative (TN) |

In this study, the binary confusion matrix for each ensemble learning model was utilized to calculate four quantitative performance metrics. Accuracy (expressed by Equation 1) represents the proportion of correctly classified samples out of the total number of samples. Precision (Equation 2) measures the proportion of positive identifications that were actually correct, focusing on the quality of positive predictions. Recall, often referred to as sensitivity (Equation 3), measures the proportion of actual positives that were correctly identified, highlighting the model’s ability to find all relevant instances. Finally, the F1-score (Equation 4) provides a balanced measure that considers both precision and recall, serving as the harmonic mean of the two.

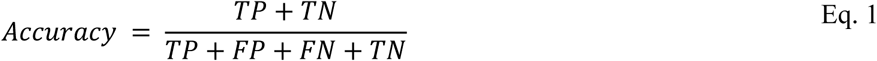

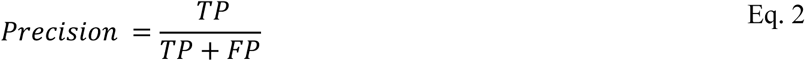

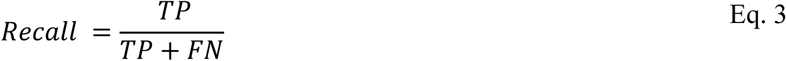

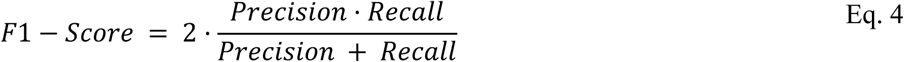

The Receiver Operating Characteristic (ROC) curve is another crucial metric for evaluating classifier performance. This curve plots the True Positive Rate (TPR) against the False Positive Rate (FPR) at various threshold settings. The Area Under the Curve (AUC) is a summary measure of the ROC curve, representing the model’s overall ability to distinguish between the two classes. An AUC value of 1.0 indicates a perfect model, while a value of 0.5 suggests a model with no discriminatory power (i.e., performance is no better than random guessing). The ROC curve provides a valuable visual tool for assessing the trade-off between sensitivity and specificity across different classification thresholds, making it an excellent way to compare the performance of multiple models.

### 2.5. Explainability Analysis with SHAP

As ML models become increasingly complex and accurate, the need for interpretability is more critical than ever. Among the various XAI techniques, SHAP has become a leading method for interpreting model predictions. Based on principles from cooperative game theory, SHAP assigns each input feature a contribution value toward a model’s output using Shapley values. It offers a consistent, model-agnostic approach that can explain predictions from any ML model, including complex ensemble and deep learning systems [83, 84].

One of SHAP’s key strengths lies in its ability to quantify both the size and direction of a feature’s impact on an individual prediction [45, 85]. This dual capability supports both global and local interpretability. Globally, SHAP generates summary plots that rank features by their overall importance across the entire dataset, helping analysts identify the most influential variables [86, 87]. Locally, SHAP provides visualizations—such as force plots and decision plots—that show how specific feature values contributed to an individual prediction relative to a baseline, offering insights into why a particular outcome was predicted [88, 89].

SHAP is especially valuable when applied to high-performing ensemble models like XGB, LightGBM, and AdaBoost, which often sacrifice transparency for predictive power [90]. By uncovering non-linear interactions, revealing hidden dependencies, and highlighting threshold effects, SHAP enhances the interpretability of these models. This makes it particularly effective for analyzing agricultural injury severity, where the relationships among driver behavior, vehicle characteristics, environmental conditions, and road/surface factors are often complex. In this study, SHAP was used to assess feature importance and interaction effects within ensemble models designed to predict agricultural injury severity. Beyond traditional performance metrics, a SHAP analysis was conducted to provide crucial insights into how and why the models made their predictions. This explainability is paramount in safety-critical domains where understanding the underlying factors is as important as achieving high accuracy. It can provide both broad policy-level insights—such as identifying key safety factors—and detailed, case-level explanations to support operational decision-making. This layered interpretability bridges the gap between prediction accuracy and practical application, offering valuable insights to first responders, agricultural safety professionals, and policymakers. In doing so, SHAP enhances the interpretability of machine learning models, transforming them from purely predictive tools into resources for both understanding and action.

## 3. Results

The performance of the ensemble ML models in predicting agricultural injury severity was comprehensively evaluated using a combination of confusion matrices, quantitative metrics, ROC curves, and SHAP-based explainability analysis. The findings indicate that while all models demonstrated strong predictive capabilities, they varied in their ability to correctly classify fatal versus non-fatal cases and in their underlying feature importance.

Table 3 presents the confusion matrices for each of the six ensemble models, offering a detailed view of their classification outcomes. A consistent and notable trend was observed across all models: a significantly higher rate of correct classifications for fatal cases compared to the non-fatal cases. For example, RF correctly predicted 1,574 fatal injuries (94%) while correctly classifying only 154 non-fatal ones (20%). This performance imbalance is directly related to the dataset’s composition, where fatal incidents constitute the majority class (68% of the records). This suggests that the models were more adept at identifying the majority class, which can sometimes come at the cost of performance on the minority class. However, models like LightGBM and HistGBRT showed a slight improvement in non-fatal classifications, but at the expense of a minor decrease in fatal prediction accuracy, a trade-off between sensitivity to the minority class and overall performance.

**Table 3.**
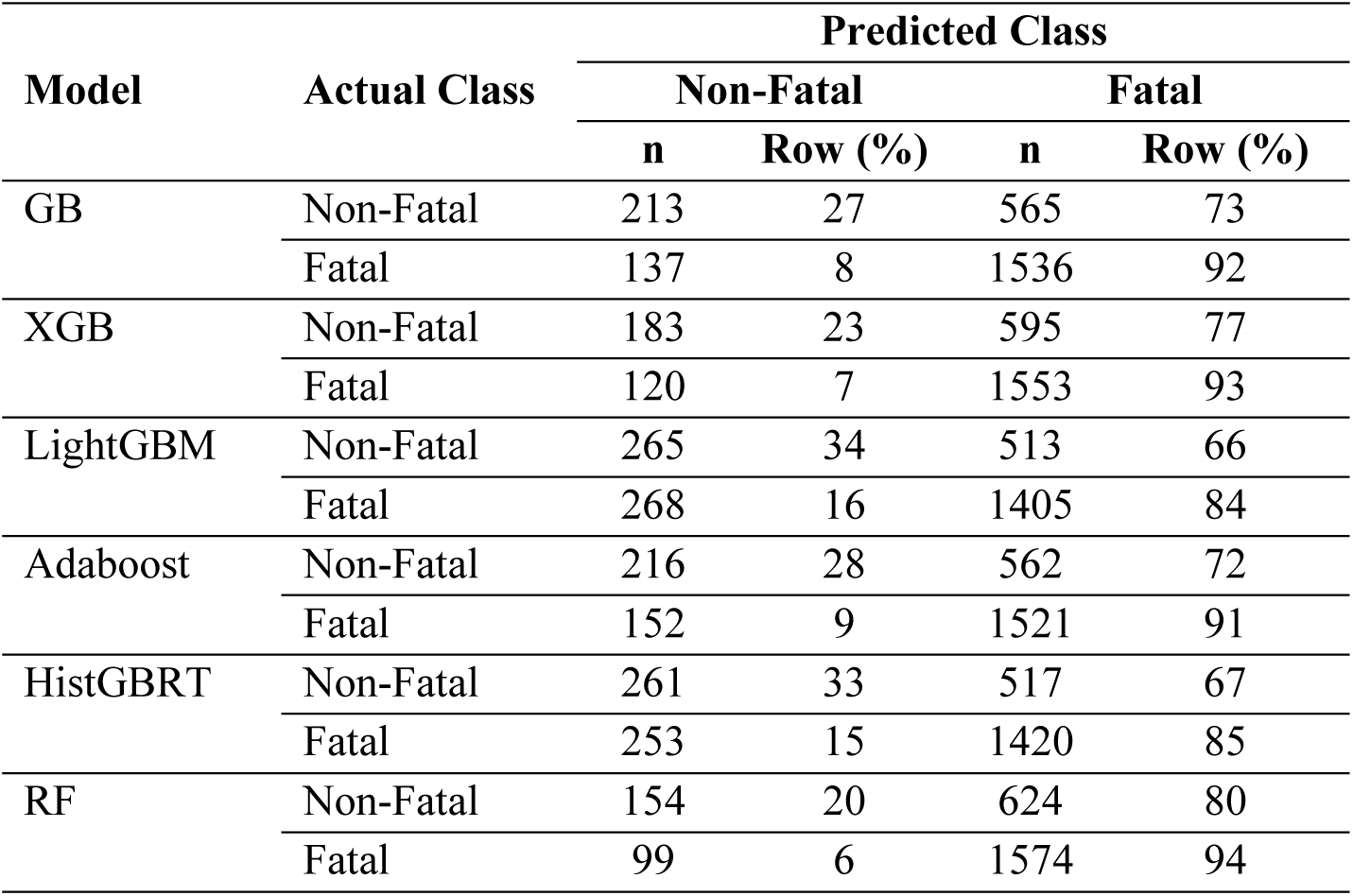
Confusion matrix for all ensemble models.

The quantitative performance metrics, summarized in Table 4, provide a more detailed comparative analysis. Most models achieved a high and consistent level of performance, with an accuracy of approximately 0.71 and a high F1-score of 0.81, indicating balanced performance between precision and recall. This consistency underscores the robustness of the ensemble approaches used for this classification task. Both RF and XGB demonstrated superior recall values (0.94 and 0.93, respectively), indicating their exceptional ability to correctly identify fatal cases. In contrast, LightGBM had the lowest overall accuracy (0.68) and F1-score (0.78), suggesting it faced more challenges in achieving a balanced performance across both classes. Despite these differences, the close metric values across most models underscore the robustness of the ensemble approaches in predicting injury severity within this dataset.

**Table 4.** Performance metrics for all ensemble models.

| **Model** | **Accuracy** | **Precision** | **Recall** | **F1-Score** |
| --- | --- | --- | --- | --- |
| GB | 0.71 | 0.73 | 0.92 | 0.81 |
| XGB | 0.71 | 0.72 | 0.93 | 0.81 |
| LightGBM | 0.68 | 0.73 | 0.84 | 0.78 |
| Adaboost | 0.71 | 0.73 | 0.91 | 0.81 |
| HistGBRT | 0.69 | 0.73 | 0.85 | 0.79 |
| RF | 0.71 | 0.72 | 0.94 | 0.81 |

The ROC curves, displayed in Figure 3, visually confirm the models’ strong performance and ability to discriminate between classes. The curves for GB, XGB, RF, and AdaBoost are positioned favorably in the upper-left quadrant of the plot, indicating high discriminative ability between the fatal and non-fatal classes. The AUC values for these models were all approximately 0.69, a strong result that is significantly better than a random guess (AUC = 0.5). While LightGBM and HistGBRT showed slightly lower AUC scores (0.67), their performance remained strong. The overall proximity of the curves highlights that all tested models effectively utilized the dataset’s features to differentiate between severity classes, with only marginal differences in their predictive capacity, making them all viable options for this type of prediction task.

**Figure 3:**
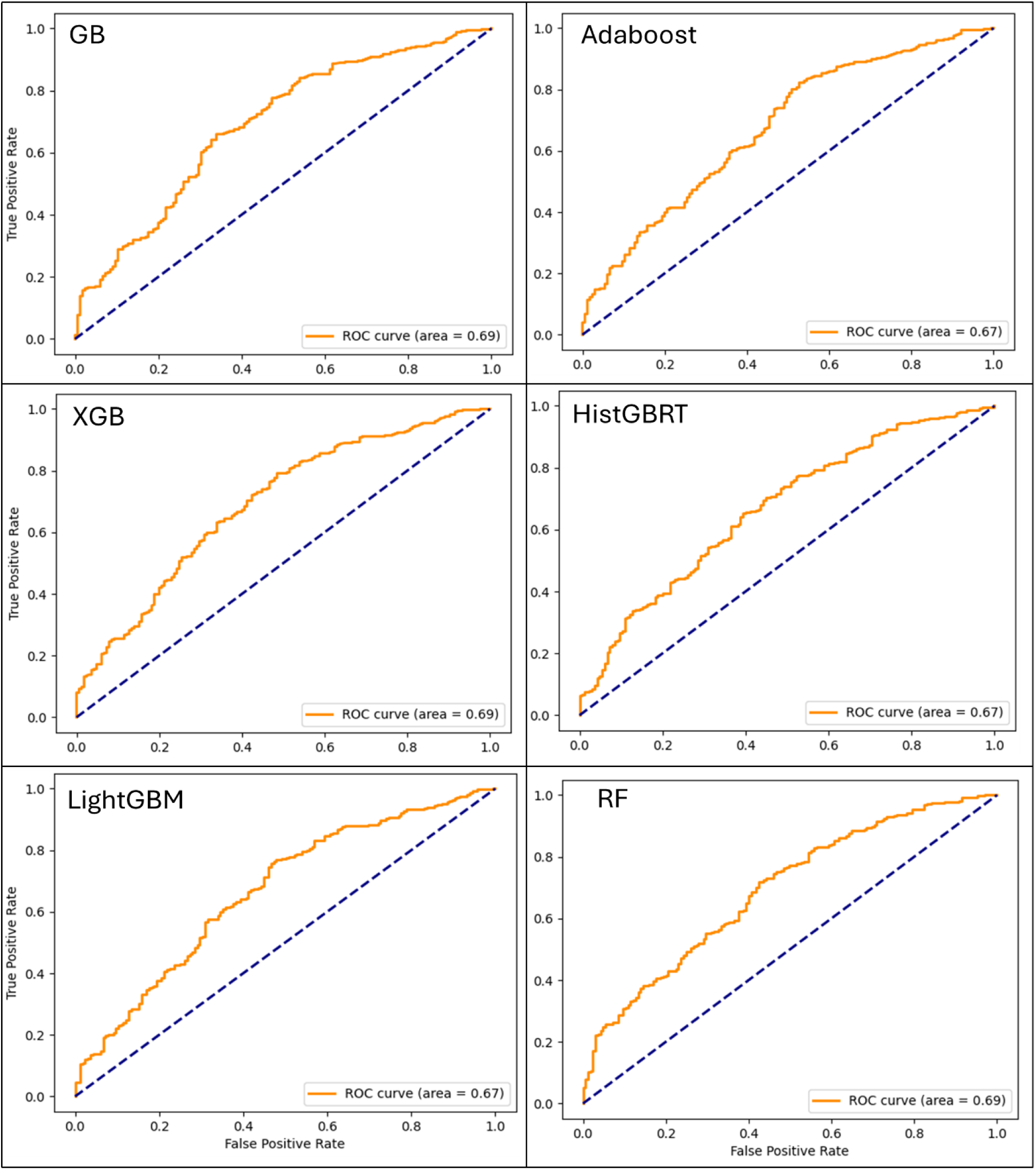
The Receiver Operating Characteristic (ROC) curves generated from Machine Learning (ML) models for injury severity classification based on data we used.

The SHAP analysis was performed at both global and local levels. The global SHAP results in Figure 4-5 provide a macro-level understanding of which features are most influential across the entire dataset. The upper panel ranks features by their mean absolute SHAP value, identifying the most influential variables. Across all models, the presence of rollover protective structures (ROPS), helmet use, victim age, and the type of injury agent emerged as the most critical factors influencing injury severity. This highlights that features related to victim protection and the incident’s direct cause are the primary drivers of prediction.

**Figure 4:**
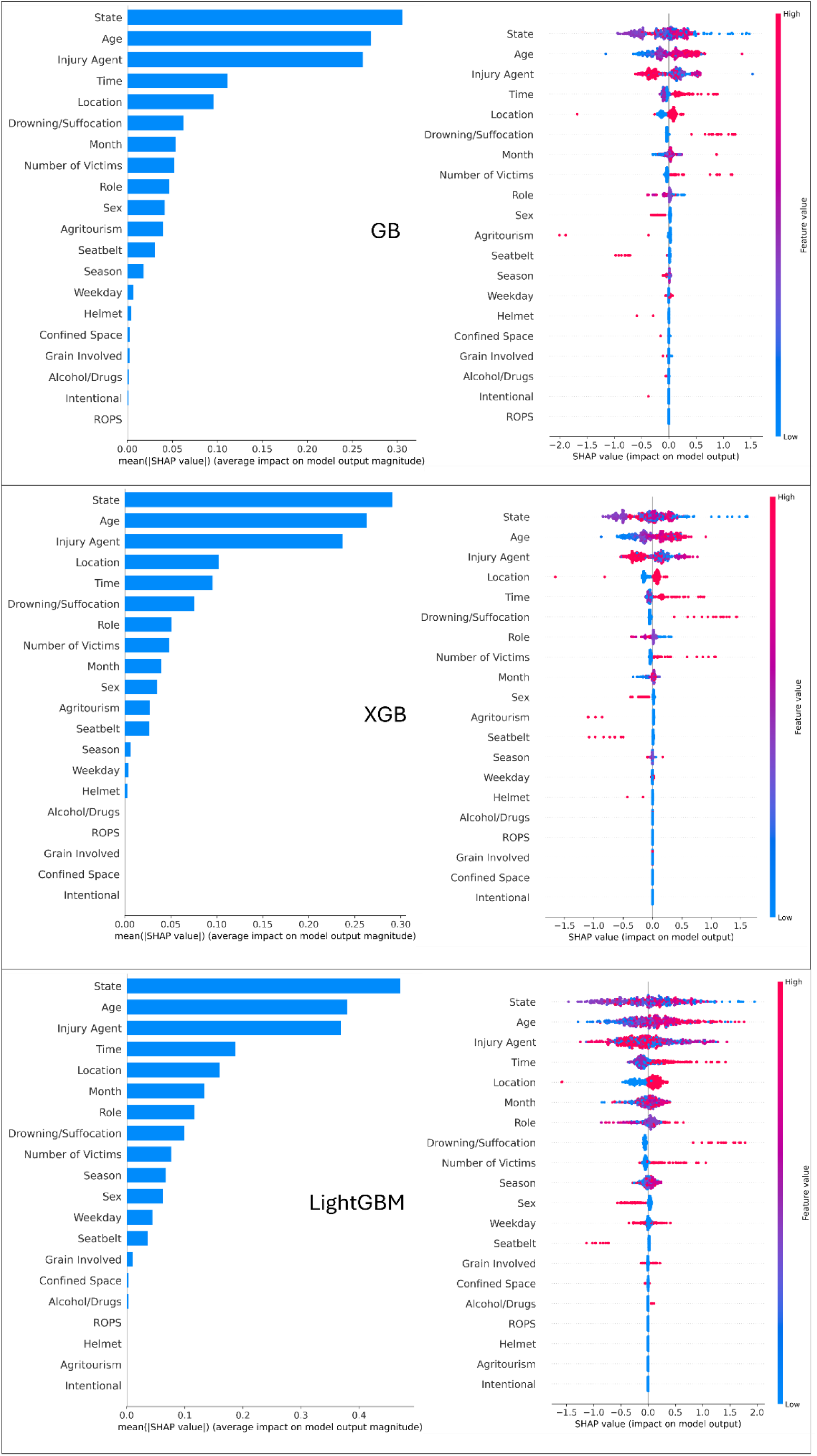
SHapley Additive exPlanations (SHAP) global interpretation for GB, XGB, and LightGBM models based on feature importance (left) and summary plot (right).

**Figure 5:**
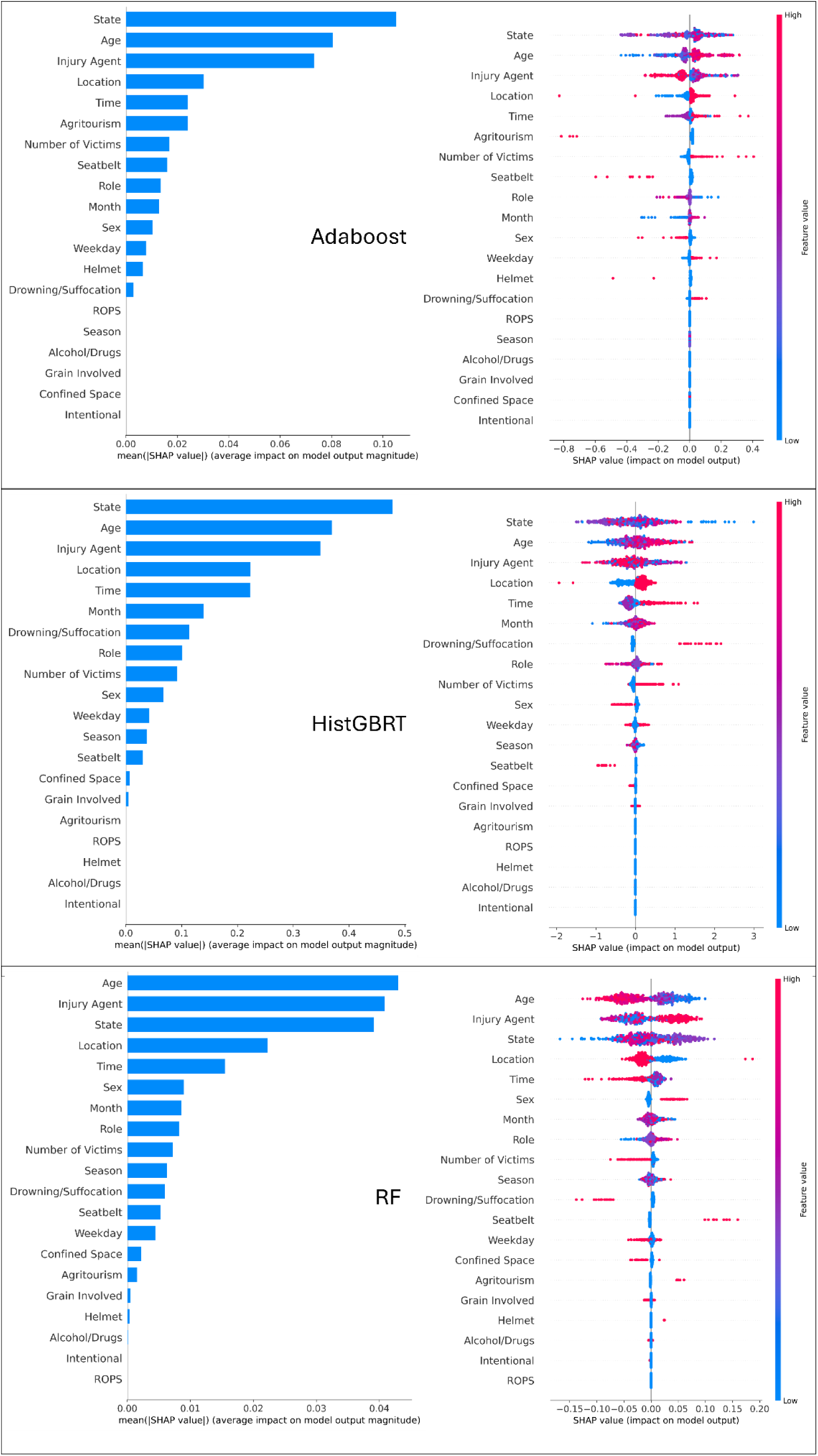
SHapley Additive exPlanations (SHAP) global interpretation for Adaboost, HistGBRT, and RF based on features importance (left) and summary plot (right).

The right panels of Figures 4 and 5 present the SHAP summary plots, which offer a deeper look at the relationship between feature values and their impact on the model’s output. Each dot represents a single prediction, with the dot’s position on the x-axis corresponding to the SHAP value (impact on model output) and its color representing the feature’s value (red for high, blue for low). The color of the dot represents the feature’s value, with red signifying a high value and blue a low value. For example, for the Age feature, the plot shows a clear trend: red dots (representing older ages) are predominantly on the positive side of the x-axis, indicating that older age tends to increase the likelihood of a fatal outcome. Conversely, for features like Helmet, red dots (indicating the presence of helmet) are clustered on the negative side, showing that this safety feature strongly decreases the likelihood of a fatal injury. These insights not only align with existing domain knowledge but also highlight practical areas for targeted safety interventions.

Figure 6 provides a vital, case-specific perspective through local SHAP interpretability plots. Unlike global analysis, which reveals general trends, these plots explain the model’s prediction for an individual incident. Each plot acts as a “force plot” for a single case, with features represented by colored bars. Red color indicates that the input variables provide stronger predictive control, while blue color indicates that the input variables contribute to weaker predictive control. The length of each bar signifies the magnitude of that feature’s influence. This level of detail is invaluable, as it demonstrates how a single variable, such as State, can have a powerful, context-dependent impact that might be obscured in a global analysis.

**Figure 6:**
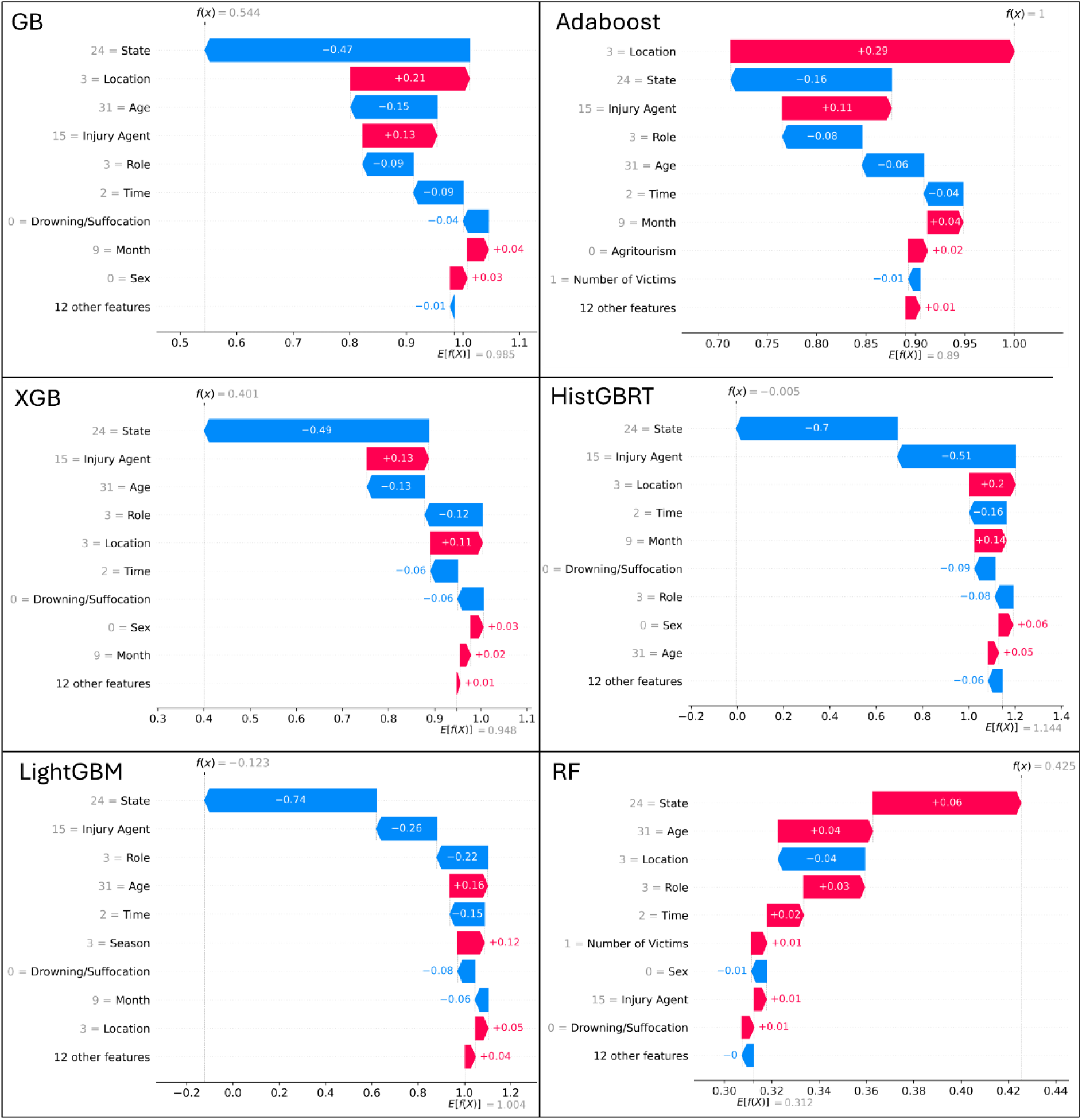
SHapley Additive exPlanations (SHAP) local interpretability plots for different Machine Learning (ML) models we use.

For instance, in one case, the specific state might be a strong contributor to fatal prediction, while in another, it might be a negligible factor. Other recurring influential features include location type, role, victim age, and injury agent, though their impact varies in magnitude and sign between models and individual cases. The input variable of Age, for example, enhanced the predictive ability for model LightGBM, RF and HistGBRT, while it decreased the predictive ability for GB, XGB and Adoboost. These local explanations reveal how the same feature can have opposite effects depending on the contextual combination of other variables, illustrating the complex interactions captured by ensemble methods. This local explainability is crucial for stakeholders like first responders, injury prevention advocates and policymakers, as it enables them to understand the specific reasons behind a predicted outcome and to design highly targeted, context-aware interventions.

## 4. Discussion

The findings of this study validate the effectiveness of advanced ensemble ML models in predicting agricultural injury severity and provide critical, interpretable insights for stakeholders. Our analysis confirms that these models can effectively leverage a unique, curated dataset to address the challenge of sparse official injury reporting in the agricultural sector. The consistent performance metrics across the models, including an average accuracy of 0.71 and an F1-score of 0.81, demonstrate the robustness and reliability of this data-driven approach.

Our model performance results align with and extend findings from previous research on crash and injury severity prediction. The superior predictive power of ensemble methods, particularly XGB and RF, echoes the conclusions of studies across various domains [12, 37, 75, 90]. These studies consistently show that ensemble techniques outperform traditional statistical and individual ML models, effectively capturing the complex, non-linear relationships inherent in injury data [30]. While our dataset’s class imbalance (68% fatal cases) posed a challenge, the robust F1-scores indicate that the models maintained a strong balance between precision and recall, consistent with findings from similar studies addressing imbalanced datasets [86, 91]. The slightly better performance of LightGBM and HistGBRT on the minority class further supports previous work on highlighting these algorithms’ efficiency and effectiveness in handling diverse data characteristics [92].

A core contribution of this work is the application of SHAP to move beyond a “black-box” approach. The global SHAP analysis, as seen in Figure 4-5, highlighted several key factors that align with existing domain knowledge in agricultural safety as well as findings from the broader traffic safety literature [46, 91]. The strong influence of safety measures like ROPS and helmet use confirms the importance of these proven interventions. Furthermore, the high importance of victim age and injury agents corroborates that demographic factors and the type of equipment involved are major determinants of agricultural injury severity. These findings are crucial for policymakers and public health professionals, as they pinpoint the most impactful areas for safety campaigns and regulatory efforts.

The local SHAP analysis presented in Figure 6 represents a novel and particularly valuable aspect of our work. Unlike a global analysis, which provides generalized insights, the local force-plots reveal the unique combination of factors driving individual predictions. For example, a victim’s geographic state or role in the incident could be a dominant factor in one case but negligible in another. This instance-specific interpretability transforms a general predictive model into a powerful diagnostic tool, a concept that is gaining traction in other fields [54, 88,89]. This level of detail is invaluable for first responders, who could use it to better understand a specific incident’s severity, while safety professionals may apply it for in-depth post-incident analyses and the development of highly targeted, context-aware training and injury prevention programs.

Our research not only demonstrates the high predictive performance of ensemble ML models for agricultural injury severity but also bridges the critical gap between accuracy and interpretability. The dataset represents a rich and multidimensional resource for modeling injury severity in agriculture. The SHAP-based findings, both global and local, provide actionable, evidence-based insights that can inform policy, guide safety interventions, and ultimately help to reduce the human and economic costs of agricultural injuries. The combination of structured incident-level data and contextual factors offers a unique opportunity to generate actionable insights for policymakers, safety professionals, and researchers in agricultural health and safety.

## 5. Conclusion

This study successfully introduced a comprehensive, data-driven framework for predicting and interpreting the severity of agricultural injuries using advanced ensemble machine learning models. By leveraging a unique dataset from AgInjuryNews, a public repository of news reports, our research directly addresses the challenge of sparse official injury data in the agricultural sector, demonstrating the viability of using publicly available news reports for large-scale injury surveillance. The comparative analysis of six different ensemble models, GB, XGB, RF, AdaBoost, LightGBM, and HistGBRT, confirmed their robust predictive capabilities, with most models achieving a high and consistent performance (accuracy of ≈0.71, F1-score of ≈0.81).

Our work’s primary contribution lies in the effective integration of XAI through SHAP to provide both global and local interpretability. The global SHAP analysis revealed that factors directly related to victim protection and incident cause, such as the presence of ROPS, helmet use, victim age, and the type of injury agent, are the most significant predictors of injury severity. These findings provide evidence-based insights that can inform broad safety campaigns and policy-making efforts. Furthermore, the local SHAP analysis offered a critical, case-specific perspective, illustrating how the influence of variables like geographic location and victim’s role can vary significantly between incidents. This level of granularity is crucial for operational decision-making and for tailoring interventions to specific high-risk scenarios.

The findings from this study have significant implications for future research and practice in agricultural safety. Our methodology provides a reproducible framework that can be applied to other domains with limited surveillance data. While the models showed strong predictive performance, future work could explore more advanced techniques to address class imbalance and further improve the prediction of less frequent injury outcomes. Additionally, the application of local explainability in real-time, in conjunction with geospatial data, could offer a powerful tool for safety organizations and first responders to proactively mitigate risks and improve incident response. In summary, our research bridges the gap between predictive accuracy and practical interpretability, providing a powerful tool to enhance safety and reduce the human and economic burden of agricultural injuries.

## Data Availability

All data produced in the present study are available upon reasonable request to the authors

## Acknowledgement

This research was funded and supported by Injury Prevention Research Center and The Iowa Initiative for Artificial Intelligence at University of Iowa.

